# Hospital saturation and risk of death without receiving mechanical ventilation in hospitalized COVID-19 patients: a city-wide analysis

**DOI:** 10.1101/2021.06.13.21258844

**Authors:** Isaac Núñez, Adrián Soto-Mota

## Abstract

**Background:** Pneumonia is the hallmark of severe COVID-19, with supplemental oxygen requirement being the main indication for hospitalization. Refractory hypoxemia in these patients requires invasive mechanical ventilation (IMV) otherwise, death is imminent. In places with a high disease burden, availability of critical care experts, beds, or resources is challenged and many patients could die without receiving them.

**Methods:** We performed a retrospective cohort study using open databases from Mexico City about suspected or confirmed COVID-19 patients, health system saturation, and deaths between May 8^th^, 2020, and January 5^th^, 2021. After building a directed acyclic graph, we performed a binary logistic regression to identify the association between proposed causal variables and dying without receiving IMV (the outcome).

**Results:** We included 33 805 hospitalized patients with suspected or confirmed COVID-19, of which 19 820 (58.6%) did not require IMV and survived, 5416 (16.1%) required and received IMV, and 8569 (25.3%) required IMV but died without receiving it. Saturation of IMV-capable beds did not increase the odds of the outcome (odds ratio 1.07, 95% confidence interval 0.94-1.22 of 90%vs50% occupancy), while general bed saturation (2, 1.86-2.14 of 90%vs50% occupancy) and IMV-capable to general bed ratio (1.64, 1.52-1.77 for a ratio of 2vs0.5) did. Private healthcare decreased the odds of the outcome (0.12, 0.08-0.17) and dyspnea increased them (1.33, 1.19-1.9).

**Conclusions:** In Mexico City, increased general hospital bed saturation and IMV-capable to general bed ratio were associated with a higher risk of dying without receiving IMV. Private healthcare was the most protective factor.

**Key messages:**

- Hospital saturation has been a central feature of public health messaging, but it is not known how outcomes relate to hospital saturation or capacity.
- In Mexico City, 90% of COVID-19 patients requiring mechanical ventilation died but less than half received it.
- Higher general bed saturation and an increased ratio of IMV-capable beds to general beds increased the probability of dying without being intubated while receiving private healthcare decreased this probability.
- Having available beds to intubate patients is possible thanks to the conversion of general beds, however, still yields suboptimal critical care.

## Introduction

Humanitarian crises compromise the availability of human and clinical resources. ^1,2^ During the COVID-19 pandemic, the scarcity of hospital beds has been one of the most impactful documented shortages. ^3,4^ Even in high-income countries, the high demand for hospitalization beds overwhelmed healthcare systems and contributed to the observed excess of deaths. ^5,6^

Since pneumonia is the hallmark of severe COVID-19 ^7^, the main indication for hospitalizing COVID-19 patients is the need for supplemental oxygen or invasive mechanical ventilation (IMV) if they develop refractory hypoxemia. ^8,9^ Therefore, multiple decisions must be made by health care providers when the demand surpasses the supply. Who should be admitted to an already saturated hospital? Of those admitted, who should be prioritized to receive mechanical ventilation? ^10^

There is no easy answer for these questions, and their answers should take into account the current hospital’s resources, the availability of other hospitals nearby, and of course, the severity of the patient’s disease. ^6,10,11^

What is more, these factors are highly dynamic and can drastically change within a few hours. A patient could arrive at a hospital that had 10 available beds that morning but filled them all by noon or could be clinically stable in the morning and quickly develop refractory hypoxemia due to pulmonary embolism, which would require a critical care bed. ^7^

Because hospital saturation was deemed inevitable in many regions ^3,12^, most countries have made efforts to increase their hospitalization capacity. ^13,14^ Mexico was not the exception and the government incremented the number of IMV-capable beds from 1800 in February 2020, to 6425 by April 2020. ^15,16^

However, getting more beds and mechanical ventilators is not enough to provide the complex and specialized care required by critically ill patients. ^17,18^ Strikingly, there are only ∼1900 registered adult or pediatric critical care specialists in Mexico ^19^ and, consequently, many patients were cared for by under-trained personnel.

Hospital saturation is one of the variables that guide Mexico’s “semáforo epidemiológico” (epidemiological traffic light), a four-stage color-coded tool that dictates the intensity of lockdown measures in a given region for the following two weeks.^20^ Both general bed and IMV-capable bed capacity are reported in daily national conferences and have never reported a 100% occupancy. ^21^ It is worth highlighting that reports that aggregate data from large regions in a country make it nearly impossible to reach 100% occupancy, even if most hospitals are completely saturated. Moreover, these regionally aggregated reports misleadingly downplay the contribution of hospital saturation to excess deaths.

Accordingly, a recent single-center study documented that of all patients admitted to a COVID-19 hospital in Mexico City that died, 45.6 % did not receive mechanical ventilation for lack of availability and 10.7% for “do not intubate/do not resuscitate” orders, suggesting that the main driver for dying without receiving IMV was indeed, hospital saturation. ^4^

Estimating the contribution of hospital saturation to mortality and its related factors is essential to guide future efforts for improving the resilience of healthcare systems. However, data from isolated hospitals or brief periods during the pandemic are suboptimal for this purpose.

In this study, we aimed to identify the factors associated with dying of COVID-19 in patients who did not receive mechanical ventilation and to estimate the net contribution of hospital saturation at a city-wide level using open data from Mexico City, which has almost 10 million habitants and has been one of the most heavily affected areas of the world during the COVID-19 pandemic. ^22^

## Methods

### Data sources

We used three open data sources: the SINAVE Mexico City COVID-19 patient database (MxCov), the death registry of Mexico City (MxDeath), and the daily reports of health system saturation (MxHcare). ^23–25^

MxCov includes anonymized clinical and socio-demographical information on all suspected COVID-19 cases that came into contact with the healthcare system (it is worth mentioning that Mexico has a complex, multi-institution healthcare system and a very large group of different private healthcare institutions). Individual identifiers, intubation status, comorbidities, site of residence, symptoms, and the healthcare system institution in which the patient was admitted were obtained from this database. MxDeath includes age, sex, cause of death as disclosed in the death certificate, a binary variable that describes if COVID-19 is mentioned as a contributing cause of death, state of residence, and the municipality of residence. Finally, MxHcare discloses information on the number and percentage of available general or IMV-capable hospital beds by healthcare system institutions.

Few dates had no daily report or had a repeated one, in these cases we used the mean of hospital saturation reported between the last and next available reports. Seven public healthcare systems (of which six are also included in MxCov) and private healthcare as a general category are included in MxHcare.

### Timeframe and patient selection

Since May 5^th^, 2020, Mexico City’s government has released daily reports about hospital availability, divided by type of bed (general and with IMV capability) and health system. The number of beds has been reported since May 8^th^, 2020, and private healthcare occupation was reported until January 5th, 2021. Thus, we included patients that were admitted into healthcare between May 8^th^, 2020, and January 5^th,^ 2021. The most recent update of the MxCov database at the time of this analysis was on April 16th, 2021. Patients were excluded if they were hospitalized outside the study period, received ambulatory treatment, or were cared for in a healthcare system with no information about occupancy. Among 55 896 suspected or confirmed COVID-19 patients included in MxCov, 33 805 were finally included in the analysis (**Figure 1**).

**FIGURE 1.**
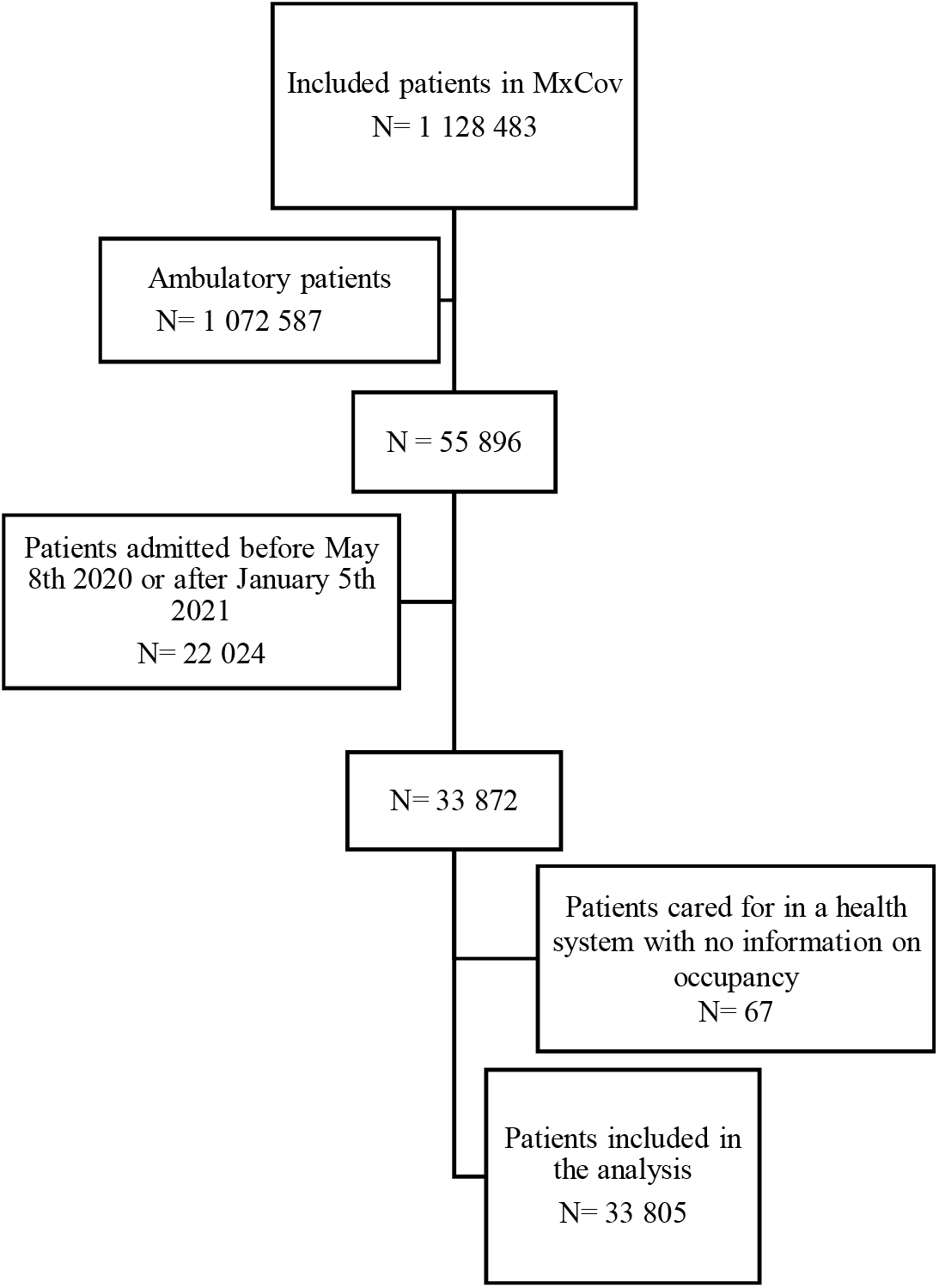
Patient selection flowchart.

### Variable definitions

COVID-19 was considered to be the cause of death in all patients of the MxCov database in whom a positive RT-PCR or antigen test was reported (confirmed COVID-19 patients). In patients who died without COVID-19 confirmation, we matched COVID-19 cases from MxCov with cases from MxDeath using: date of death, age, sex, state of residence, municipality of residence, and hospital as the place of death. Since MxDeath has individual identifiers for each registered death, it is possible to identify those patients who had more than one matching factor without having other patients with the same characteristics. If a death certificate reported confirmed or suspected COVID-19 as one of the causes of death, we considered it to be the cause of death.

Finally, three kinds of patients were identified: those who were hospitalized did not require IMV (and survived), those who required and received IMV, and those who died without receiving IMV.

### Statistical analysis

The MxCov database includes patients with suspected or confirmed COVID-19 and, during the first stage, we performed the analysis using all hospitalized patients. We built a directed acyclic graph (DAG) using the ‘DAGitty’ online software version 3.0 to identify which variables could be related to patients dying because of not being intubated even though they needed it (the outcome). ^26,27^ Before building the DAG, we hypothesized that three variables were the main direct causes of this outcome: lack of bed availability, patient’s desire not to be intubated, and medical futility because of disease severity (**Figure 2**). None of these variables were directly observed.

**FIGURE 2.**
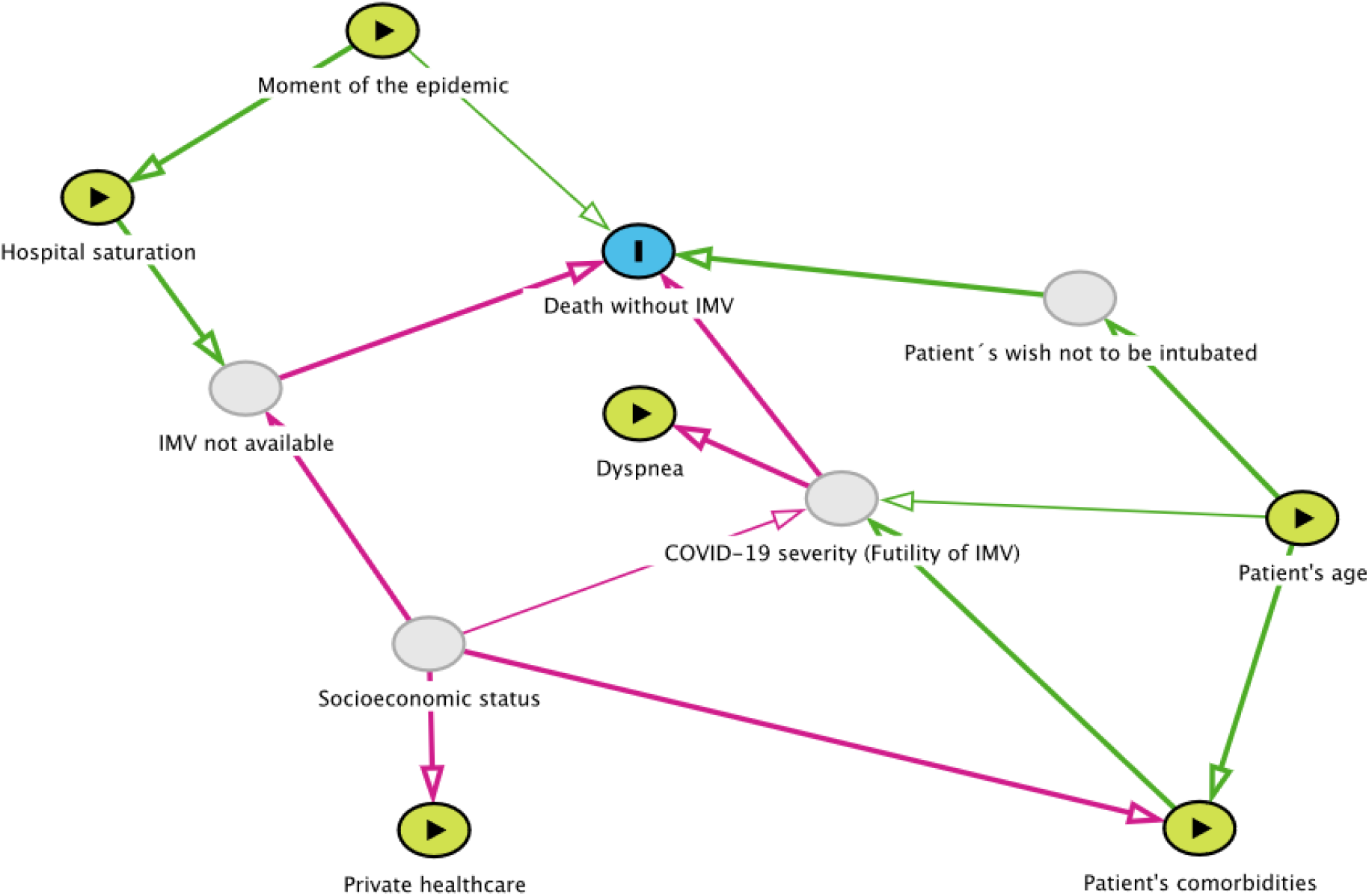
Directed Acyclic Graph (DAG) of hypothesized causal variables on death without receiving invasive mechanical ventilation (IMV). The blue node is the outcome, green nodes are observed exposures, and gray nodes are unobserved exposures. Hospital saturation was replaced by IMV-capable to general bed ratio in an additional adjusted logistic regression model.

Based on our DAG, we can estimate; the effect of the moment of the pandemic (represented by epidemiological week) on the outcome, the effect of hospital saturation (considering IMV-capable beds and general beds saturation in separate models) by controlling for the epidemiological week, the effect age by controlling for patient comorbidities, the effect of patient comorbidities by controlling for age and partially for socioeconomic status using private healthcare as a surrogate variable, the effect of private healthcare (as a surrogate for socioeconomic status) by controlling for patient comorbidities, and the effect of dyspnea (as a surrogate for disease severity) by controlling for patient age and partially for socioeconomic status using private healthcare as a surrogate variable.

We calculated the ratio of available IMV-capable beds and general hospital beds as an indicator of increased hospital-bed conversion and placed it in place of hospital saturation in an adjusted model (**Figure 2**).

We used descriptive statistics for analyzing the clinical and socio-demographical variables of patients that were hospitalized and did not require IMV, those that were hospitalized and received IMV, and those that were hospitalized and died without receiving IMV. We assumed those patients that received IMV and those that died without receiving it both required IMV.

We performed binary logistic regression models to obtain odds ratio and corresponding 95% confidence intervals to evaluate the association of epidemiological week, age, presence of diabetes, obesity, hypertension, hospital saturation, epidemiological week (a surrogate for the moment of the pandemic), and private healthcare (a surrogate for socioeconomic status) with dying of COVID-19 without being intubated against being intubated. Age, hospital saturation, and IMV-general bed ratio were included in the models as continuous variables using cubic splines with three nodes. ^28^

All analyses were conducted with R software version 4.0.0.

## Results

### Clinical and socio-demographical characteristics

Patient distribution was as follows: 19 820 (58.6%) did not require IMV and survived, 5416 (16.1%) required and received IMV, and 8569 (25.3%) required IMV but died without receiving it. Of those who required and received IMV 4129 (76.2%) died, resulting in a mortality of 90.7% (12 698 people out of the 13 985 patients that required mechanical ventilation). Clinical and socio-demographical characteristics are shown in **Table 1**.

**TABLE 1.** Clinical and sociodemographic characteristics of hospitalized COVID-19 patients according to the outcome.

| **Variable** | **Did not require**<br>**IMV (survived)** | **Received IMV** | **Died without**<br>**receiving IMV** |
| --- | --- | --- | --- |
| **Number of patients**<br>**(n= 33 805)** | 19 820 (58.6) | 5416 (16.1) | 8569 (25.3) |
| **Age (median, IQR)** | 54 (40-65) | 61 (51-70) | 66 (56-75) |
| **Sex** | - | - | - |
| **-Female** | 8805 (44.4) | 1893 (35) | 3105 (36.2) |
| **-Male** | 11 015 (55.6) | 3523 (65) | 5464 (63.8) |
| **Positive antigen or**<br>**RT-PCR test** | 9380 (47.3) | 3325 (61.4) | 5317 (62) |
| **Deaths** | 0 (0) | 4129 (76.2) | 8569 (100) |
| **Comorbidities** | - | - | - |
| **-Obesity** | 3632 (18.3) | 1239 (22.9) | 1491 (17.4) |
| **-Hypertension** | 5458 (27.5) | 2010 (37.1) | 3487 (40.7) |
| **-Diabetes** | 4681 (23.6) | 1764 (32.6) | 2959 (34.5) |
| **-Heart disease** | 960 (4.8) | 273 (5) | 473 (5.5) |
| **Presence of**<br>**dyspnea** | 12 893 (65.1) | 4425 (81.7) | 6895 (80.5) |
| **Private healthcare** | 1712 (8.6) | 325 (6) | 105 (1.2) |

Continuous variables are expressed as the median and interquartile range (IQR). Counts are expressed as the total and percentage. IMV: invasive mechanical ventilation, RT-PCR: real- time-polymerase chain reaction.

### Regression analysis

Results of the logistic regression analysis are shown in **Table 2**. Results for continuous variables and epidemiological week are also shown in **Figure 3**. Being admitted at a later epidemiological week was associated with increased odds of death without IMV (OR 1.44; 95% CI 1.34-1.54 comparing week 54 with week 37) while an early admission conferred decreased odds (0.68, 95% CI 0.64-0.73 comparing week 9 with week 37). A higher percentage of IMV-capable bed occupation was not associated with increased odds of death without IMV (OR 1.07; 95% CI 0.94-1.22 comparing 90% with 50% saturation). Lower saturation of IMV-capable beds was associated with increased odds of dying without IMV (OR 1.31; 95% CI 1.23-1.4 comparing 30% with 50% saturation). Conversely, a higher percentage of general bed occupation was associated with increased odds of death without IMV (OR 2; 95% CI 1.86-2.14 comparing 90% with 50% saturation). An elevated ratio of IMV-capable to general beds increased the odds of not receiving IMV (OR 1.64, 95% CI 1.52-1.77 comparing 2 with 0.5) but a decreased ratio dropped the odds (OR 0.47, 95% CI 0.42-0.52 comparing 0.1 with 0.5).

**TABLE 2.** Logistic regression analysis of possible causal variables and the outcome of death without receiving invasive mechanical ventilation.

| **Variable** | **OR (95% CI)** |
| --- | --- |
| **Epidemiological week** | - |
| 19 | 0.68 (0.64-0.73) |
| 27 | 0.81 (0.78-0.84) |
| 37 | Reference |
| 47 | 1.24 (1.19-1.29) |
| 54 | 1.44 (1.34-1.54) |
| **Percentage occupation of IMV-capable beds** | - |
| 30% | 1.31 (1.23-1.4) |
| 50% | Reference |
| 70% | 0.97 (0.94-1.01) |
| 90% | 1.07 (0.94-1.22) |
| **Percentage occupation of general beds** | - |
| 30% | 1.05 (0.89-1.22) |
| 50% | Reference |
| 70% | 1.2 (1.13-1.27) |
| 90% | 2 (1.86-2.14) |
| **IMV-capable to general bed ratio** | - |
| 0.1 | 0.47 (0.42-0.52) |
| 0.3 | 0.69 (0.65-0.73) |
| 0.5 | Reference |
| 0.8 | 1.4 (1.33-1.48) |
| 1 | 1.56 (1.45-1.67) |
| 2 | 1.64 (1.52-1.77) |
| **Private healthcare** | 0.12 (0.08-0.17) |
| **Age** | - |
| 20 | 0.18 (0.15-0.23) |
| 30 | 0.33 (0.28-0.37) |
| 50 | Reference |
| 70 | 2.28 (2.15-2.42) |
| 80 | 3.13 (2.88-3.4) |
| **Diabetes** | 1.18 (1.06-1.3) |
| **Hypertension** | 1.09 (0.98-1.2) |
| **Obesity** | 0.89 (0.79-1) |
| **Dyspnea** | 1.33 (1.19-1.5) |

Details of covariate adjustment for individual models are provided in the methods section.
OR: Odds ratio; CI: Confidence interval; IMV: Invasive mechanical ventilation

**FIGURE 3.**
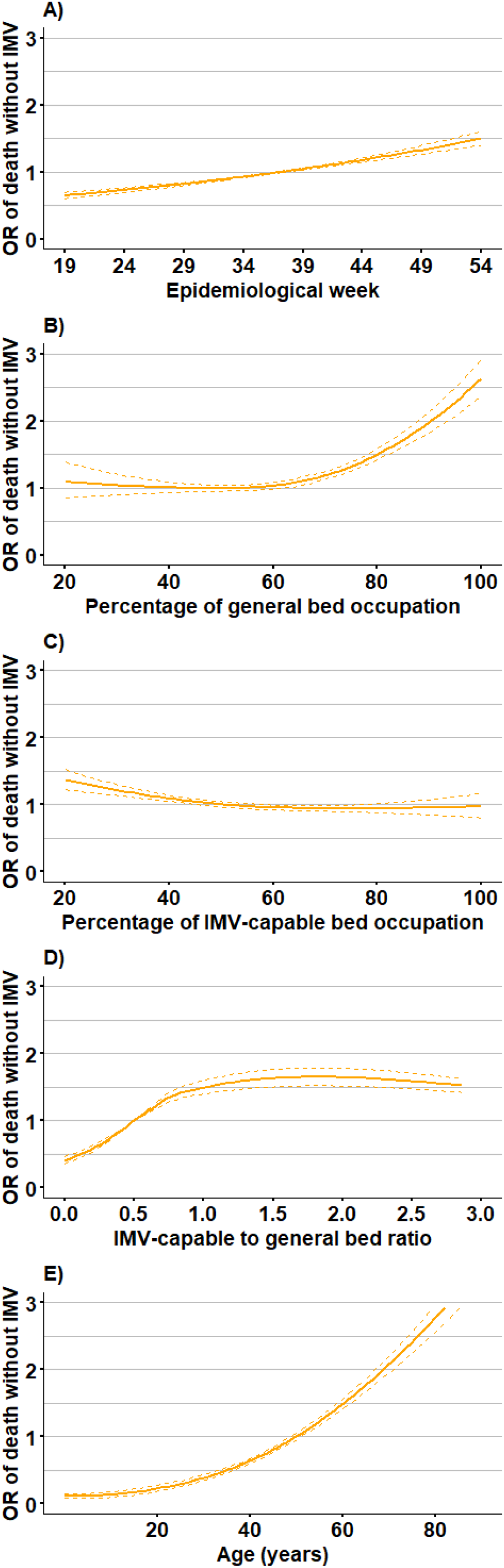
Association between proposed causal variables and dying without receiving invasive mechanical ventilation. Variables included are A) epidemiologic week, B) the percentage of general bed occupation, C) percentage of invasive mechanical ventilation-capable bed occupation, D) invasive mechanical ventilation-capable to general bed ratio, and E) age. OR: Odds ratio; IMV: invasive mechanical ventilation. Shown are adjusted OR (except epidemiological week, which was not adjusted, details on adjustments are disclosed in the methods section) with corresponding 95% confidence intervals.

Private healthcare substantially decreased the odds of the outcome (OR 0.12, 95% CI 0.08-0.17), while having dyspnea increased the odds (OR 1.33, 95% CI 1.19-1.5). Among comorbidities, diabetes and hypertension were associated with higher odds (OR 1.18 and 1.09), and obesity with lower odds (OR 0.89).

## Discussion

To our knowledge, this is the first study trying to estimate the attributable proportion of deaths to hospital saturation in Mexico. Additionally, evaluated which factors are associated with dying of COVID-19 without having received IMV.

It is paramount to make a difference between patients who did not desire to receive IMV, from those who did not receive it because their clinicians considered it futile, and, finally, from those who wanted to receive it, were considered eligible by their healthcare providers and had no access to it.

A patient’s desire to be or not intubated is a complex choice. Literature is scarce, but available studies report that age and pre-existing diseases are associated with this. ^29,30^ Futility refers to a very low probability of survival and substantial morbidity, thus a lack of benefit from invasive maneuvers. While this is extremely complex in itself, deciding who gets a bed or a ventilator among people in whom critical care normally would not be considered to be futile. ^10^ Normally, futility would be the thing that a medical team would consider to determine if a patient benefits or not from critical care. ^29^

Interestingly, a recent survey among physicians put good prognosis, lower age, and having caregiving responsibilities among the main criteria to account for when choosing who gets the last ICU bed. ^31^ When proposed with several methods to choose who gets the last bed, most chose to use a pre-established set of rules by the health department or chose to defer the decision to the senior physician. ^31^

Medical futility is extremely complex to decide and can only be accounted for by having a direct conversation with the medical team that treated a given patient. Variables that could be related to this are acute disease severity, pre-existing comorbidities, age, and short expected survival with any therapeutic maneuver. We used dyspnea, for example, as a surrogate for disease severity. The increased odds of outcome seen in these patients could be due to increased disease severity (futility) but this is only a possible symptom of severe disease and some patients present with “happy hypoxemia”, in which low oxygenation is asymptomatic. ^32^

To explain the counterintuitive lower odds we observed of dying without being intubated precisely when IMV-capable beds were most saturated, it is worth highlighting that a higher IMV-capable-beds to general-beds-ratio suggest that more general beds were “converted” into IMV-capable beds to deal with the high demand of IMV. Therefore, the number of patients receiving IMV increased, and consequently, the proportion of patients who died without it declined. In summary, the higher demand for IMV saturated the general bed’s pool and turned IMV-bed saturation into an inaccurate pandemic severity indicator.

This is further supported by the extremely high mortality of 90% among patients that required intubation, as improvised and suboptimal care became more likely because the number of healthcare providers with critical care training remained constant. For context, a recent international study in African high care or intensive care units reported a “high mortality” of 48.9%. ^33^

The lower odds of dying without receiving mechanical ventilation we observed in those cared for in private healthcare institutions is consistent with the worse general outcomes seen in people with lower socioeconomic status in Mexico and other Latin American countries. ^34,35^ This underlines the need for increased support for those in a vulnerable position.

Furthermore, the outcomes difference between different epidemiological weeks is consistent with a previous analysis in which the place of death (hospital or home) differed according to the date of death, with domiciliary deaths increasing from 8% to 21% by the end of 2020. ^36^

## Strengths and limitations

Among the limitations of our study are the use of repurposed data and the lack of individual center data. Even if we matched hospital saturation with their respective health system affiliation, individual hospitals could have many different levels of occupation.

We cannot estimate the direct effect of clinical futility and socioeconomic status, which are important variables that could influence the decision of not offering or receiving IMV. However, a previous study conducted in Mexico City showed a low proportion of patients voluntarily refusing IMV, and it is reasonable to expect this was the case in other hospitals. ^4^ Additionally, the fact that the median age of those who died without IMV was low (66 years old) and that their comorbidities frequency and distribution was similar in the other groups, suggests it is unlikely that clinical futility was a frequent reason for not offering critical care or that a large proportion of patients voluntarily refused to receive IMV.

On the other hand, our study also has several strengths. We used open data from a large metropolitan area that is updated regularly. Additionally, drew a DAG before analyzing or determining which were the most probable relationships between variables and what was reasonable to expect from our analysis. Finally, because of how our data was curated, we could reasonably ascertain the cause of death in most cases.

## Conclusion

Our study suggests that the higher demand for IMV forced the conversion of general hospital beds and their saturation. The high mortality we found is consistent with this because hospital beds can be quickly increased but specialized personnel takes time to train. While we cannot ascertain the wishes of every patient, it is highly unlikely that most people voluntarily refused to be intubated, our data suggest that hospital saturation was indeed, a cause of death in these patients.

Regional aggregates of hospital saturation are inaccurate indicators and should not be emphasized while communicating the pandemic status to the population because they sub estimate the high contribution of hospital saturation to mortality.

## Data Availability

All utilized code and data will be made freely available with the final version of the article.

## Ethics approval

This study does not require ethics board approval because it used publicly available anonymized data, with no interaction between the researchers and individuals.

## Data availability

All utilized data is freely available in respective government webpages and all used code will be openly available with the final version of the article.

## Acknowledgments

We would like to thank medical residents, nursery personnel, and other healthcare workers. Your work has been invaluable. Also, we thank scientists around the world who have devoted themselves to continue working and supporting science communication.

